# Human Infection with Avian Influenza A(H7N7) Virus, Taiwan, 2026

**DOI:** 10.64898/2026.10.09.26365013

**Authors:** Yao-Tsun Li, Yun-Tong Lin, Bao-Shan Chen, Chuan-Yi Kuo, Hsiang-Yi Huang, Hui-Ying Ko, Pei-Yu Huang, Gavin JD Smith, Yi-Ling Lin, Ming-Tsan Liu

## Abstract

Low pathogenic avian influenza A(H7N7) virus detected in a poultry worker in Taiwan, 2026 was related to viruses circulating in Asian wild birds and possessed PB2 E627K, a mammalian adaptation marker. Enhanced surveillance for H7 viruses in poultry and rapid characterization of human cases are essential to assess pandemic potential.

**Article Summary Line:** Identification of a wild bird-origin avian influenza A(H7N7) virus with a PB2 E627K substitution in a poultry worker highlights the need for enhanced surveillance.

## Main

Avian influenza A(H7) viruses have caused sporadic zoonotic infections in humans, with most documented cases epidemiologically linked to direct poultry exposure or contaminated poultry-associated environments (1). Among H7 subtypes, human infections with highly pathogenic avian influenza (HPAI) H7N7 viruses were reported in the Netherlands in 2003 and Italy in 2013, resulting in 89 and 3 human cases, respectively (1,2). Cases in these major outbreaks occurred among poultry workers or persons involved in culling operations. Amino acid residues associated with human adaptation or increased pathogenicity have emerged in H7 virus genomes during animal or human outbreaks (1), highlighting the importance of closely monitoring these viruses for public health risk assessment. We report a human infection with low pathogenic avian influenza (LPAI) H7N7 virus in Taiwan, confirmed by quantitative PCR (qPCR), serological assays, and whole-genome sequencing.

A male duck farm worker was admitted to a hospital with fever on a day in 2026, two days after onset of mild respiratory symptoms (3). The patient had underlying chronic medical conditions and subsequently developed pneumonia, for which antiviral treatment was administered. After clinical improvement and negative follow-up influenza test results, he was discharged 14 days after illness onset. No avian influenza virus was reported to have been detected in the duck farms visited by the patient (3).

Using qPCR targeting the matrix (M) gene, we detected influenza A virus in the patient’s throat swab collected 5 days after illness onset, with a cycle threshold (Ct) value of 38.4. The sample was negative for influenza B virus and for other subtype-specific qPCR tests targeting the hemagglutinin (HA) genes of H1, H3, H5, and H7 viruses. A sputum sample collected 7 days after illness onset showed a stronger signal for influenza A virus (Ct 32.8), with a weak H7-specific signal detected at a Ct value of 40.0, whereas results for other influenza A subtypes tested negative. All close contacts tested negative for influenza virus by the same qPCR approach. Attempts to recover virus by cell culture were unsuccessful, probably because of low viral load or antiviral treatment.

Serological testing by hemagglutination inhibition (HI) assay and enzyme-linked immunosorbent assay (ELISA) further supported H7 subtype infection (Supplementary Methods). HI titers against an H7N7 virus, A/duck/Taiwan/A45/2003, increased twofold from 7 to 12 days after illness onset, and fourfold from 12 to 20 days, reaching a titer of 1:160 (Figure 1A). In contrast, HI titers against an H5N2 virus, A/duck/Taiwan/21080001/2021, which previously circulated in Taiwan (4), remained unchanged at 1:20 across all three sampling time points. Similar findings were observed by ELISA. Antibody levels against the H7 HA protein (A/Shanghai/02/2013) increased continuously from 7 to 20 days after illness onset, and ELISA absorbance values against H7 HA were at least 10-fold higher than those against H5 HA protein (A/dairy cow/New Mexico/A240920343-93/2024) in each serum sample (Figure 1B). None of the paired serum samples collected from contacts 14 days apart showed a >2-fold increase in antibody titers by either HI or ELISA (data not shown).

**Figure 1.**
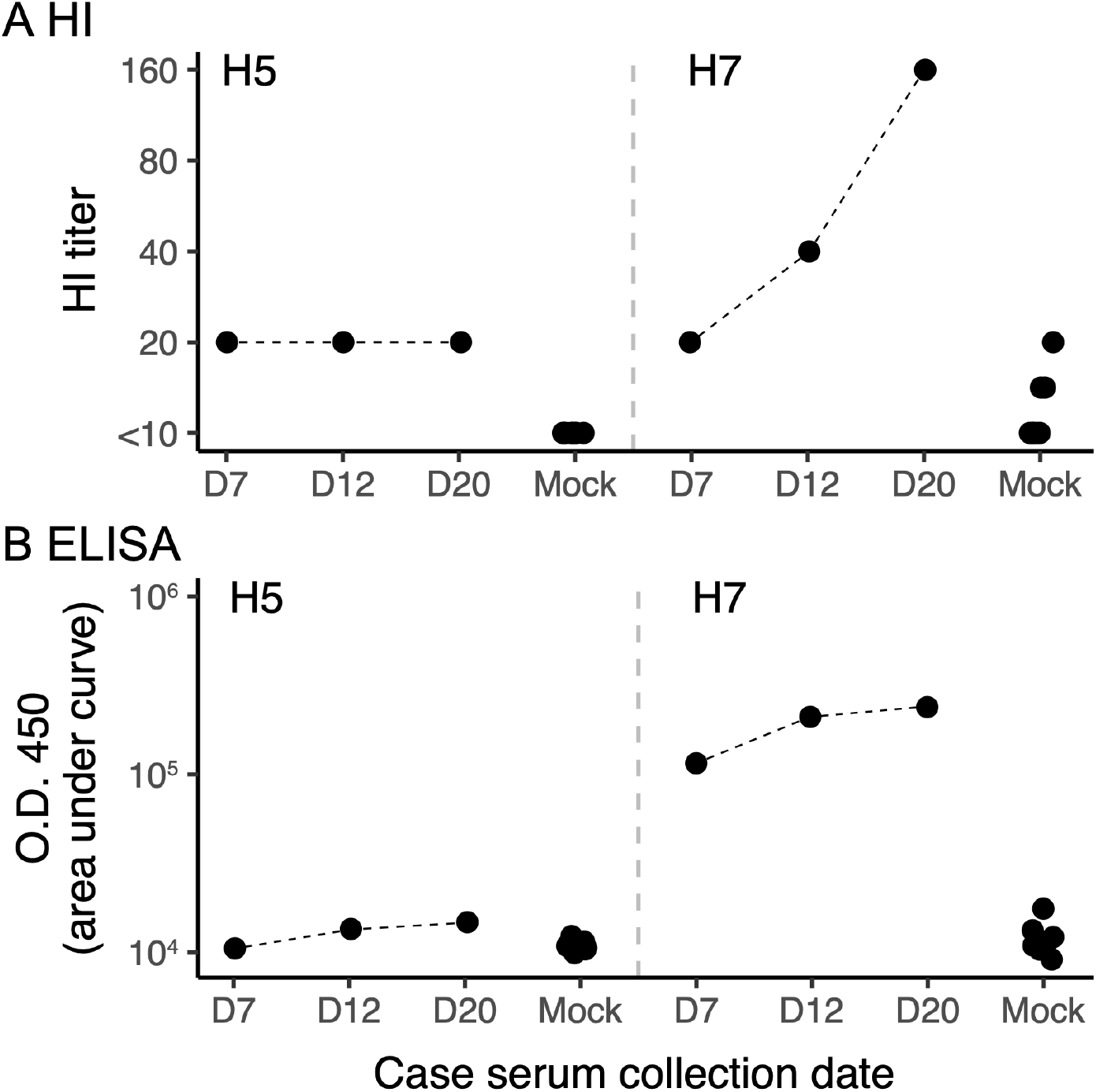
Antibody responses against H5 and H7 influenza viruses in the patient with influenza A(H7N7) virus infection. Antibody titers were measured by (A) HI assay and (B) ELISA using serum samples collected from the patient on days 7, 12, and 20 after illness onset. Serum samples from persons previously reported for suspected zoonotic influenza cases but who tested negative were included as mock controls (n=8).

To further characterize the virus, we performed whole-genome sequencing using viral RNA amplified by influenza genome-specific PCR and sequenced on a nanopore platform (Supplementary Methods). The sequencing library prepared from the sputum sample collected 7 days after illness onset showed substantial coverage when reads were mapped to an H7N7 virus, A/Mallard/South Korea/KNU2021-49/2021, with median depth for each gene segment ranging from 272 to 2266 reads. Consensus sequences generated from the library identified the virus as subtype H7N7. The HA and NA segments shared >98% nucleotide identity with publicly available H7 and N7 genes (Supplementary Table).

Phylogenetic analyses of the H7N7 virus, hereafter named A/Taiwan/1/2026, showed that all gene segments were closely related to viruses isolated from wild birds or domestic waterfowl in Asia during 2023-2025 (Figure 2; Supplementary Figure), indicating that the virus originated from the Eurasian avian influenza gene pool. The virus did not share an identical gene constellation with viruses available in public databases. Although the HA and M genes clustered with an A/environment/Kagoshima/KU-24-D10/2024(H7N7)-like virus isolated in Japan, the other gene segments were most closely related to different source viruses. Across the genome, A/Taiwan/1/2026 is distantly related to previously detected viruses from Taiwan and to reported human H7 viruses from China, Europe, and North America (Figure 2; Supplementary Figure). These findings suggest a recent spillover from the Eurasian gene pool to a human, distinct from previous major H7N7 human infections in which viruses had adapted to or circulated in terrestrial poultry before transmission to humans (5). Based on the patient’s occupation background, domestic ducks at the workplace may have served as an intermediate source of infection after introduction of the virus into Taiwan by migratory birds. However, direct exposure to migratory birds or their feces could not be excluded.

**Figure 2.**
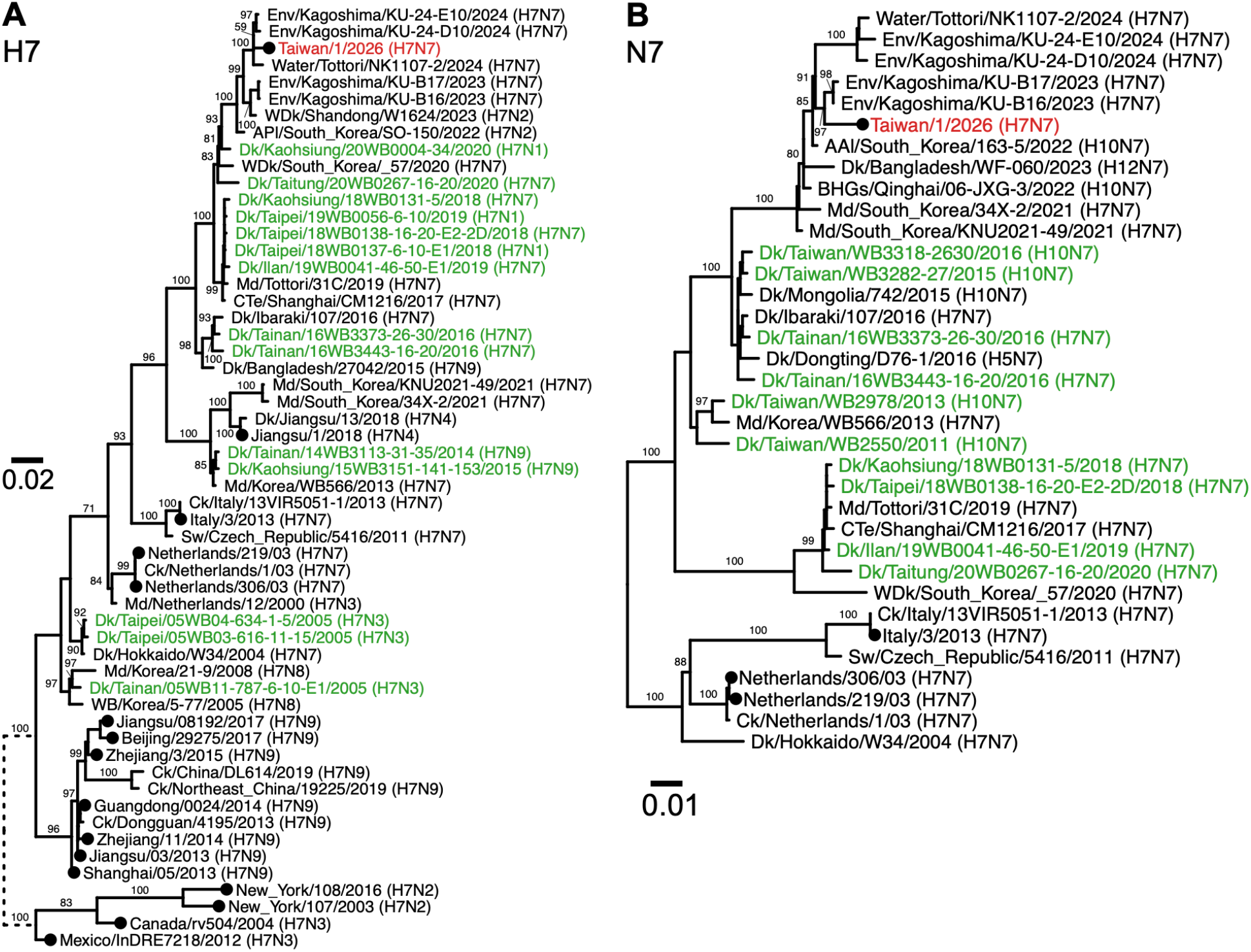
Maximum likelihood phylogenies of the HA(H7) and NA(N7) genes of the H7N7 virus identified in this study. The virus sequenced from the patient sample is labeled in red, and viruses isolated in Taiwan are labeled in green. Black circles at the terminal nodes indicate viruses from previously reported human infections. Selected bootstrap support values are shown. The long branches at the tree root are shortened and shown as dashed branches. AAl, Anser albifrons; APl, Anas platyrhynchos; BHGs, bar-headed goose; Ck, chicken; CTe, common teal; Dk, duck; Env, environment; Md, mallard; Sw, swan; WB, wild bird; WDk, wild duck.

We examined molecular signatures of A/Taiwan/1/2026 to assess its pathogenicity, receptor binding, mammalian adaptation, and antiviral susceptibility (Table). The virus possessed two basic amino acids at the HA cleavage site (PEIPKGR|G) and was therefore inferred to be low pathogenic (6). The virus likely retained α2,3-linked sialic acid receptor-binding preference, given the absence of key HA residue changes associated with human-type receptor binding, including G186V and Q226L, which have been reported in H7N9 human viruses (6). An E627K substitution was identified in PB2, whereas other mammalian adaptive markers, including K526R and D701N, were not observed. E627K is a well-studied substitution that enhances replication of avian influenza viruses in mammalian hosts (7), and is frequently identified in viruses involving human infections (1). Because the virus was sequenced during a later phase of infection (day 7) and closely related avian viruses available in databases possessed 627E in PB2 (Table), PB2 E627K in A/Taiwan/1/2026 might have emerged during replication in the patient. However, its presence in the infecting avian virus cannot be excluded. We detected neither PA I38T associated with reduced susceptibility to endonuclease inhibitors nor the NA substitutions E119V, R152W, E276D, R292K, and D293N associated with reduced susceptibility to neuraminidase inhibitors (8). These findings suggest that A/Taiwan/1/2026 has no known molecular markers of reduced antiviral susceptibility.

**Table.**
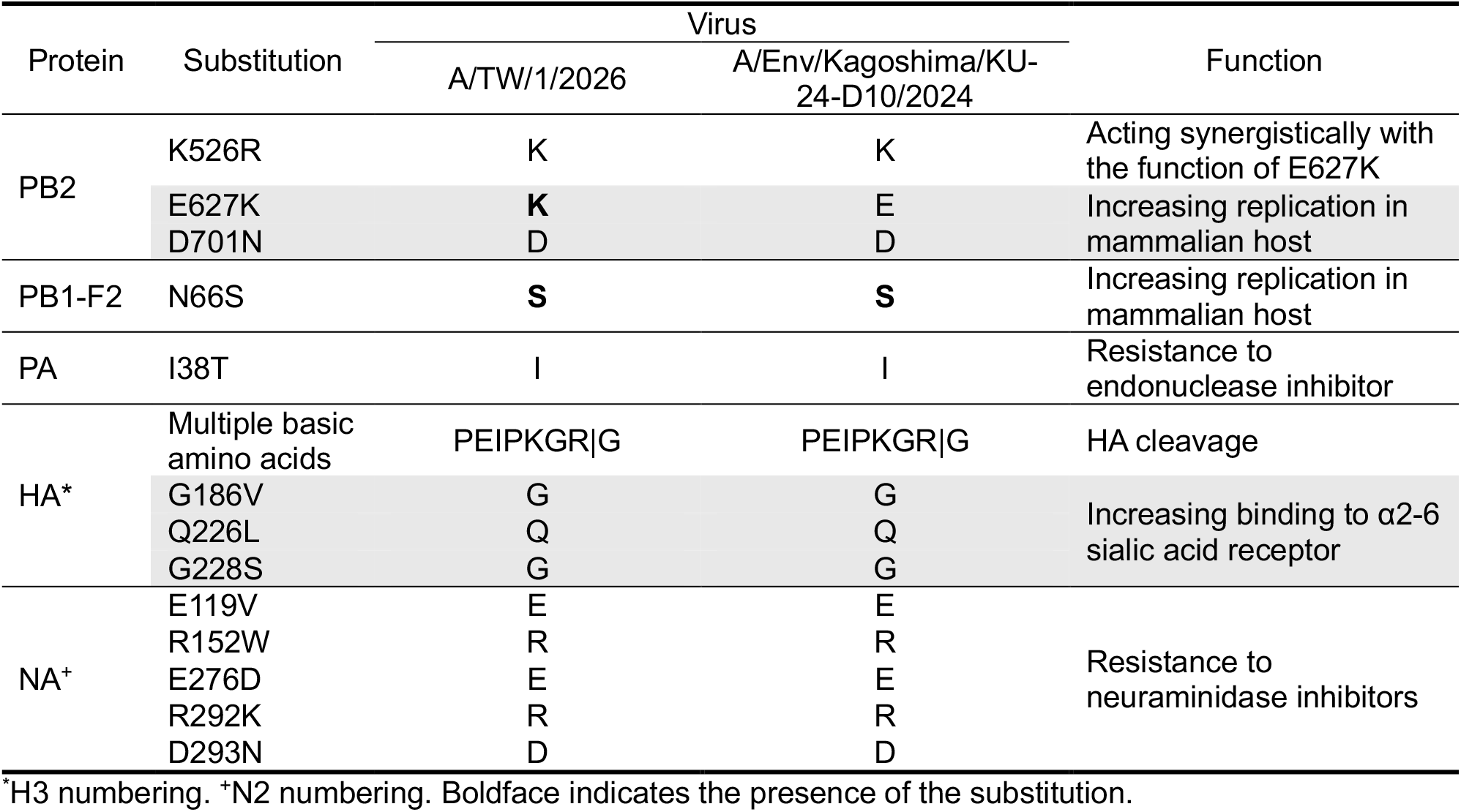
Molecular signatures of A/Taiwan/1/2026 (H7N7) and a closely related H7N7 virus.

## Conclusions

We confirmed human infection with LPAI H7N7 virus in Taiwan, supported by genomic and serological evidence. Phylogenetic analyses show that A/Taiwan/1/2026 originated from the Eurasian gene pool harbored by wild birds and domestic waterfowl, although the exact route of transmission to the patient remains unclear. Functional amino acid profiling indicates the virus is an LPAI virus and likely retains avian-type receptor binding specificity. The presence of PB2 627K in A/Taiwan/1/2026 suggests enhanced replication potential in mammalian hosts.

Although the risk for efficient human-to-human transmission of A/Taiwan/1/2026 and related viruses from the Eurasian gene pool requires further characterization, this case highlights a potential risk of direct or indirect transmission of wild bird-origin avian influenza viruses to humans, especially among persons at the animal-human interface. As Taiwan is located at the middle of East Asian and Australasian Flyway (9), this case reveals a potential surveillance gap in detecting avian influenza viruses moving from northern Asia into Taiwan and reaching humans with bird exposure. Strengthened surveillance among wild birds and poultry, together with rapid data sharing, will be critical for clarifying virus dissemination pathways.

## Data Availability

The original data presented in this study can be found at https://github.com/yaotli/taiwan_h7n7. Genome sequences produced in this study are available from the NCBI GenBank database (accession numbers: PZ812265-PZ812272).

## Acknowledgments

We acknowledge the health care practitioners in the notifiable disease network for sample collection. We gratefully acknowledge Dr. An-Suei Yang and Dr. Chung-Ming Yu of the Genomics Research Center, Academia Sinica, and Dr. Chun-Che Liao of the Biomedical Translation Research Center, Academia Sinica, for providing the recombinant H5 and H7 hemagglutinin proteins used in this study. We acknowledge the authors, originating laboratories, and submitting entities responsible for providing the sequences obtained from GISAID. We would like to thank National Core Facility for Biopharmaceuticals (NCFB, NSTC 115-2740-B-492-001) and National Center for High-performance Computing (NCHC) of National Institutes of Applied Research (NIAR) of Taiwan for providing computational and storage resources.

## Supplementary Methods

### Real-time PCR

Viral RNA was extracted from throat swabs or sputum specimens using the TANBead Maelstrom 4800 platform with the corresponding nucleic acid extraction kit (Taiwan Advanced Nanotech). One-step real-time reverse transcription PCR was performed using the LightCycler Multiplex RNA Virus Master Kit (Roche) with primers modified from World Health Organization (WHO) protocols (1). Assays were used to detect influenza A and B viruses and to subtype seasonal influenza A(H1N1)pdm09 and A(H3N2) viruses. An H5-specific primer set originally developed for clade 2.3.4.4 H5Nx viruses (1) and an H7-specific primer set developed for influenza H7N9 viruses (2) were used to detect H5 and H7 viruses, respectively.

### HI

Serum antibody titers against H5 and H7 viruses were measured by hemagglutination inhibition (HI) assay as previously described (3). The representative strains A/duck/Taiwan/A45/2003 (H7N7) and A/duck/Taiwan/21080001/2021 (H5N2) were kindly provided by the Veterinary Research Institute, Taiwan.

### ELISA

Serum H5- and H7-specific IgG antibodies were quantified by indirect enzyme-linked immunosorbent assay (ELISA) to corroborate the HI results. 96-well microplates were coated overnight at 4°C with recombinant H5 hemagglutinin (HA) protein derived from H5N1 virus A/dairy cow/New Mexico/A240920343-93/2024 (GISAID accession no. EPI3257776), or recombinant H7 HA protein derived from H7N9 virus A/Shanghai/02/2013 (NCBI accession no. NC_026425) at 0.2 μg/mL in carbonate coating buffer (Thermo Fisher Scientific). Plates were washed with phosphate-buffered saline containing 0.1% Tween-20 (PBST) and blocked with 5% skim milk at 4°C overnight. Human serum samples were serially diluted four-fold in blocking buffer from 1:10 to 1:163,840 and incubated in antigen-coated wells for 1 h at room temperature. After washing with PBST, bound antibodies were detected with horseradish peroxidase (HRP)– conjugated goat anti-human IgG antibody (Sigma-Aldrich, catalog no. A0170) for 1 h at room temperature. Plates were washed again with PBST, developed with 3,3’,5,5’-tetramethylbenzidine (TMB) substrate (Sigma-Aldrich) for 10 min at room temperature, and the reaction was stopped with stop solution. Absorbance was measured at 450 nm using a microplate reader. Antibody responses were quantified by calculating the area under the curve (AUC) from ELISA titration curves in GraphPad Prism version 11.

### Nanopore and bioinformatic analysis

The viral genome was amplified by PCR using influenza-specific primers (4). PCR product was subjected to end repair and dA-tailing using the NEBNext Ultra II End Repair/dA-Tailing Module (New England Biolabs). Barcodes were ligated using the Native Barcoding Kit (EXP-NBD114.96, Oxford Nanopore Technologies) and Blunt/TA Ligase Master Mix (New England Biolabs). Barcoded products were pooled and ligated to Native Adapters (Oxford Nanopore Technologies) using the NEBNext Quick Ligation Module (New England Biolabs). The final library was loaded onto an R10.4.1 flow cell (FLO-MIN114, Oxford Nanopore Technologies) and sequenced on a MinION device according to the manufacturer’s instructions. Basecalling and demultiplexing of pod5 files were performed with Dorado v1.3.0 (https://github.com/nanoporetech/dorado). Reads with a quality score <8 were discarded. Consensus sequences of the influenza genome were generated using a bioinformatic workflow described previously (5). Read coverage across each gene segment was assessed with bedtools v2.31.1 (6).

### Phylogenetic analyses

Genome sequences of influenza A viruses related to A/Taiwan/1/2026 and H7 viruses associated with human infection were downloaded from GISAID (https://gisaid.org) on April 20, 2026 (7). Sequences were aligned with MAFFT v7.520 (8) and trimmed to coding regions. Maximum likelihood phylogenetic trees were inferred using IQ-TREE v2.2.0 (9) with a GTR+G model and 10,000 ultrafast bootstrap replicates.

**Supplementary Figure S1.**
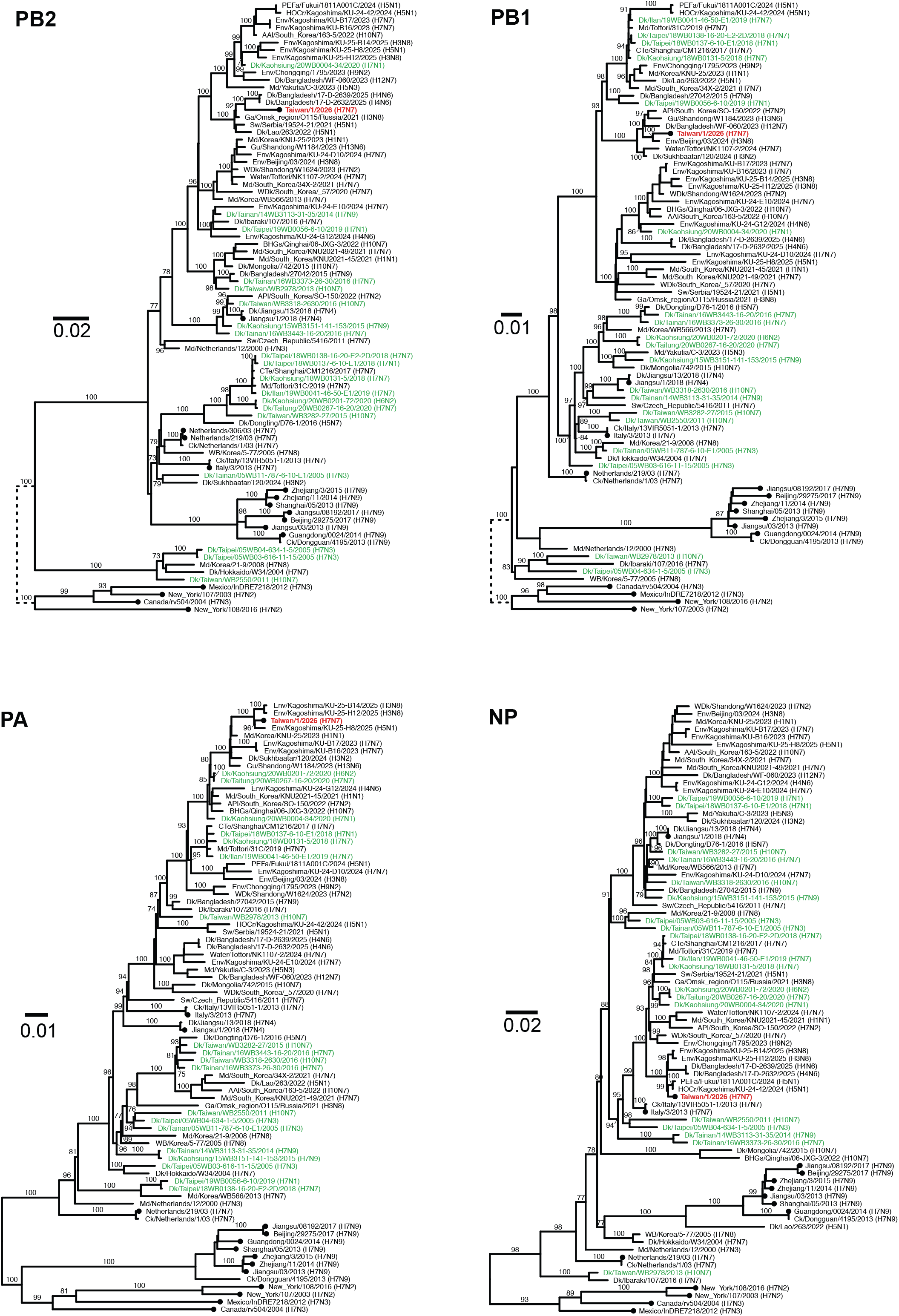
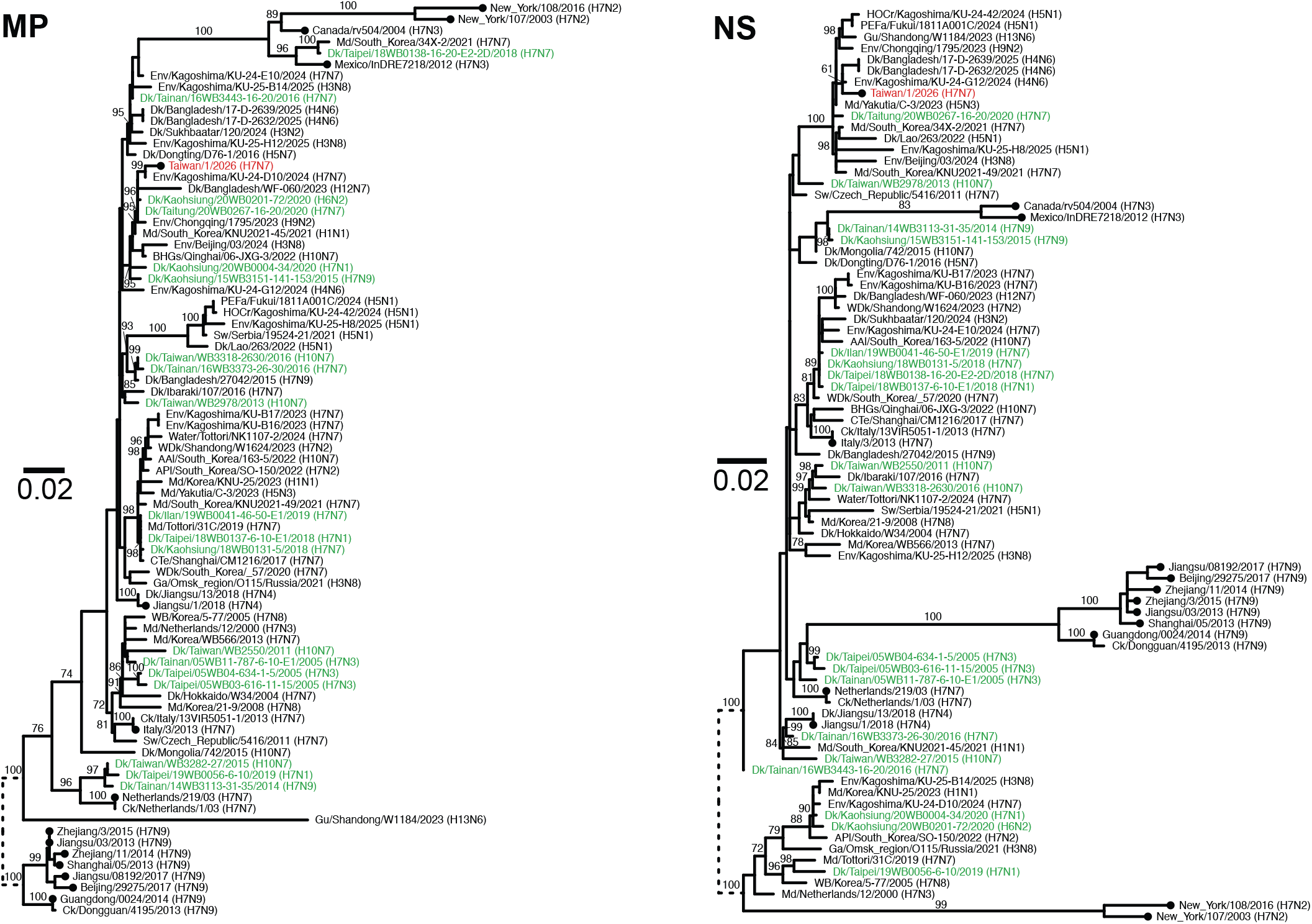
Maximum likelihood phylogenies of the internal genes of the H7N7 virus identified in this study. The virus sequenced from the patient sample is labeled in red, and viruses isolated in Taiwan are labeled in green. Black circles at the terminal nodes indicate viruses from previously reported human infections. Selected bootstrap support values are shown. The long branches at the tree root are shortened and shown as dashed branches. AAl, Anser albifrons; APl, Anas platyrhynchos; BHGs, bar-headed goose; Ck, chicken; CTe, common teal; Dk, duck; Env, environment; Ga, garganey; Gu, gull; HOCr, hooded crane; Md, mallard; PEFa, peregrine falcon; Sw, swan; WB, wild bird; WDk, wild duck.

**Supplementary Table S1.** Nucleotide sequences of A/Taiwan/1/2026 (H7N7) compared with related sequences from GISAID.

| Gene segment | Virus with highest identity | Identity* (%) | Accession no. |
| --- | --- | --- | --- |
| PB2 | A/garganey/Omsk_region/O115/Russia/2021 (H3N8) | 97 (2221/2280) | EPI2886636 |
| PB1 | A/environment/Beijing/03/2024 (H3N8) | 98 (2249/2275) | EPI4488153 |
| PA | A/environment/Kagoshima/KU-25-H8/2025 (H5N1) | 99 (2134/2151) | EPI4899090 |
| HA | A/environment/Kagoshima/KU-24-E10/2024 (H7N7) | 98 (1661/1683) | EPI4463970 |
| NP | A/environment/Kagoshima/KU-24-H12/2024 (H4N6) | 99 (1488/1497) | EPI4357871 |
| NA | A/environment/Kagoshima/KU-B16/2023 (H7N7) | 98 (1395/1416) | EPI3052742 |
| M | A/environment/Kagoshima/KU-24-D10/2024 (H7N7) | 99 (976/982) | EPI4332757 |
| NS | A/environment/Kagoshima/KU-24-G12/2024 (H4N6) | 99 (831/838) | EPI4323141 |

\* Including only coding regions.

## References

1. Li YT, Linster M, Mendenhall IH, Su YCF, Smith GJD. Avian influenza viruses in humans: lessons from past outbreaks. Br Med Bull. 2019 Dec 1;132(1):81–95. doi:10.1093/bmb/ldz036

2. Puzelli S, Rossini G, Facchini M, Vaccari G, Di Trani L, Di Martino A, et al. Human Infection with Highly Pathogenic A(H7N7) Avian Influenza Virus, Italy, 2013. Emerg Infect Dis. 2014 Oct;20(10):1745–9. doi:10.3201/eid2010.140512

3. Taiwan CDC. Statement on Taiwan’s First Locally Acquired Human Case of Novel Influenza A(H7) [Internet]. 2026 Apr 2. Available from: https://www.cdc.gov.tw/Bulletin/Detail/bZE85LXA9ZGdCvEJKZe6Cg?typeid=9

4. Chen LH, Lin YJ, Liu YP, Li WC, Tu YC, Hu SC, et al. Distinct infectivity, pathogenicity, and adaptive evolution of clade 2.3.4.4 H5N2 avian influenza viruses in chickens in Taiwan. Poultry Science. 2026 Jul 1;105(7):106913. doi:10.1016/j.psj.2026.106913

5. Fouchier RAM, Schneeberger PM, Rozendaal FW, Broekman JM, Kemink SAG, Munster V, et al. Avian influenza A virus (H7N7) associated with human conjunctivitis and a fatal case of acute respiratory distress syndrome. Proceedings of the National Academy of Sciences. 2004 Feb 3;101(5):1356–61. doi:10.1073/pnas.0308352100

6. Xiong X, Martin SR, Haire LF, Wharton SA, Daniels RS, Bennett MS, et al. Receptor binding by an H7N9 influenza virus from humans. Nature. 2013 Jul;499(7459):496–9. doi:10.1038/nature12372

7. Long JS, Mistry B, Haslam SM, Barclay WS. Host and viral determinants of influenza A virus species specificity. Nat Rev Microbiol. 2019 Feb;17(2):67–81. doi:10.1038/s41579-018-0115-z

8. Song MS, Marathe BM, Kumar G, Wong SS, Rubrum A, Zanin M, et al. Unique Determinants of Neuraminidase Inhibitor Resistance among N3, N7, and N9 Avian Influenza Viruses. Journal of Virology. 2015 Oct 2;89(21):10891–900. doi:10.1128/jvi.01514-15

9. Li YT, Chen CC, Chang AM, Chao DY, Smith GJD. Co-circulation of both low and highly pathogenic avian influenza H5 viruses in current poultry epidemics in Taiwan. Virus Evol. 2020 Jan 1;6(1):veaa037. doi:10.1093/ve/veaa037

## References

1. WHO. WHO information for the molecular detection of influenza viruses. 2021. Available from: https://cdn.who.int/media/docs/default-source/influenza/molecular-detention-of-influenza-viruses/protocols_influenza_virus_detection_feb_2021.pdf

2. WHO. Real-time RT-PCR Protocol for the Detection of Avian Influenza A(H7N9) Virus. 2013. Available from: https://cdn.who.int/media/docs/default-source/influenza/global-influenza-surveillance-and-response-system/related-documents/cnic_realtime_rt_pcr_protocol_a_h7n9.pdf

3. Yang JR, Kuo CY, Huang HY, Wu FT, Huang YL, Cheng CY, et al. Characterization of Influenza A (H7N9) Viruses Isolated from Human Cases Imported into Taiwan. PLOS ONE. 2015 Mar 6;10(3):e0119792. doi:10.1371/journal.pone.0119792

4. Zhou B, Donnelly ME, Scholes DT, St. George K, Hatta M, Kawaoka Y, et al. Single-Reaction Genomic Amplification Accelerates Sequencing and Vaccine Production for Classical and Swine Origin Human Influenza A Viruses. Journal of Virology. 2009 Oct;83(19):10309–13. doi:10.1128/jvi.01109-09

5. Perlas A, Reska T, Croville G, Tarrés-Freixas F, Guérin JL, Majó N, et al. Improvements in RNA and DNA nanopore sequencing allow for rapid genetic characterization of avian influenza. Virus Evol. 2025 Jan 1;11(1):veaf010. doi:10.1093/ve/veaf010

6. Quinlan AR, Hall IM. BEDTools: a flexible suite of utilities for comparing genomic features. Bioinformatics. 2010 Mar 1;26(6):841–2. doi:10.1093/bioinformatics/btq033

7. Shu Y, McCauley J. GISAID: Global initiative on sharing all influenza data – from vision to reality. Eurosurveillance. 2017 Mar 30;22(13):30494. doi:10.2807/1560-7917.ES.2017.22.13.30494

8. Katoh K, Standley DM. MAFFT Multiple Sequence Alignment Software Version 7: Improvements in Performance and Usability. Mol Biol Evol. 2013 Apr 1;30(4):772–80. doi:10.1093/molbev/mst010

9. Minh BQ, Schmidt HA, Chernomor O, Schrempf D, Woodhams MD, von Haeseler A, et al. IQ-TREE 2: New Models and Efficient Methods for Phylogenetic Inference in the Genomic Era. Mol Biol Evol. 2020 May 1;37(5):1530–4. doi:10.1093/molbev/msaa015

